# COPD and social distancing in the UK

**DOI:** 10.1101/2022.02.08.22270657

**Authors:** Mark David Walker, Hakan Lane

## Abstract

**Background:** Those with Chronic Obstructive Pulmonary Disease (COPD) were at a higher risk of experiencing severe illness in the event of contracting COVID-19. Did they therefore act more cautiously?

**Aim:** The aim was to determine whether the condition of COPD incurred significant change in social distancing behavior compared to the general public.

**Design and Setting:** Data was used from the Imperial ‘COVID-19 Behavioural Tracker’, which details results of regular public surveying on attitudes surrounding COVID-19 guidance.

**Methods:** Responses by U.K. participants to twenty questions reflecting willingness to adhere to social distancing guidance were compared in those reporting COPD and non-COPD status.

**Results:** Those with COPD stated a significantly greater willingness to wear face masks during early stages of pandemic. There was greater reluctance to go out and go shopping. There was no apparent or significant difference in willingness to use public transport, suggesting that this was an unavoidable necessity for all. The relationship between level of adherence and COVID-19 case numbers was weak both for those of both COPD and non-COPD status.

**Discussion:** These results suggest that those with COPD were more cautious and followed guidance more willingly. Advice provided by GP’s and healthcare professionals is likely to be beneficial in guiding patient behaviour.

## Introduction

The SARS-COV2 virus spread across the World from early 2020 and has caused wide upheaval of many societies. In Great Britain, it has been estimated that over 7 million citízens (11 % of the population) were infected and 135000 lives were lost, as of September 18, 2021. It has caused a very hard strain on the health care system and the economy (1).

The Government and local authorities have adopted a wide range of policy measures to halt and mitigate the propagation of the virus, including three national “lockdowns”, periods where many businesses were ordered to close and people ordered to work from their residences when possible. There has also been issued a number of behavioural guidelines and social distancing advice through official channels. Some of the most important advice were:

- Wear face mask
- Avoid gatherings
- Wash hands regularly
- Cover mouth when sneezing
- Reduce intimate contacts

The respiratory nature of symptoms reported by those initially experiencing COVID-19 infection means that those with Chronic Obstructive Pulmonary Disease (COPD) are at greater risk of experiencing severe illness or of mortality should they contract the condition (2, 3). Thus, advice was given that those with COPD should be particularly strenuous in their efforts to reduce the risk of contracting COVID-19 infection (4). Stringent guidance was given to those with such pre-existing health care issues to reduce social contacts and restrict daily social activities to reduce the chances of infection (5). It has been mentioned that individuals with COPD felt a higher need to self-isolate, which to some degree led to heightened emotions of loneliness (6). One study found extremely high prevalence of terror, fear and apprehension during a lockdown period (7). It has been identified that in particular the experiences of living through respiratory problems gave COPD patients an elevated level of anxiety, as lung impairment was frequently reported in COVID positive people (8).

From early 2020, the Institute of Global Health Innovation at Imperial College in London has organized regular weekly surveying of the general public on social distancing habits and personal perceptions relating to COVID-19, freely available on the ‘COVID-19 Behaviour Tracker’ (9). Respondents are questioned about 13 specific pre-existing health issues, one of which is COPD.

The paper aims to examine whether COPD patients in the UK adhered to more stringent social distance measures than the general public during the pandemic. The share answering either “Always” or “Frequently” to a set of 20 questions related to compliance with reducing contacts, increased hygiene and avoiding intimacy are analysed and hypothesis tests used to see which saw a difference between the cohorts. A subset of the questions were chosen to generate a timeline over compliance through the studied period.

## Methods

Survey data between 1 April 2020 to 16 June 2021 was used, comprising 43 rounds of surveying conducted at approximately 1 to 3 week intervals. There are 20 questions related to social distancing as presented in Table 1.

**Table 1.** Hypothesis tests social distancing.

|  |
| --- |
| **How often have you...** |
| Worn a face mask outside your home? |
| Washed hands with soap and water? |
| Used hand sanitizer? |
| Covered your nose and mouth when sneezing or coughing? |
| Avoided contact with people who have symptoms or you think may have been exposed to the coronavirus? |
| Avoided going out in general? |
| Avoided going to hospital or other healthcare settings? |
| Avoided taking public transport? |
| Avoided working outside your home? |
| Avoided letting your children go to school/ university? |
| Avoided having guests to your home? |
| Avoided small social gatherings (not more than 2 people)? |
| Avoided medium-sized social gatherings (between 3 and 10 people)? |
| Avoided large-sized social gatherings (more than 10 people)? |
| Avoided crowded areas? |
| Avoided going to shops? |
| Slept in separate bedrooms at home, when normally you would share a bedroom? |
| Eaten separately at home, when normally you would eat a meal with others |
| Cleaned frequently touched surfaces in the home (e.g. doorknobs, toilets, taps)? |
| Avoided touching objects in public (e.g. elevator buttons or doors)? |

The proportion of answers in the two highest categories (“Always” and “Frequently”) were derived for each answer for the COPD and the Healthy group, and z test for proportions carried out. The level of compliance on four of these questions, considered to be indicative of general social activity, were studied in time series:

1. Willingness to wear a face mask or covering outside the home (Question: i12_health_1),
2. Avoided going out in general (Question: i12_health_6).
3. Avoided taking public transport (Question: i12_health_8).
4. Avoided going to shops (Question: i12_health_16).

For these questions, a z test was conducted once for each monthly period.

### Baseline Data

A total of 43450 people were surveyed. Age were distributed between 18 and 96 years with a median of 48, and interquartile range of 34 – 63. There were 52,5 % females and 47,5 % males. The shares of employment status are shown in Figure 1.

**Figure 1.**
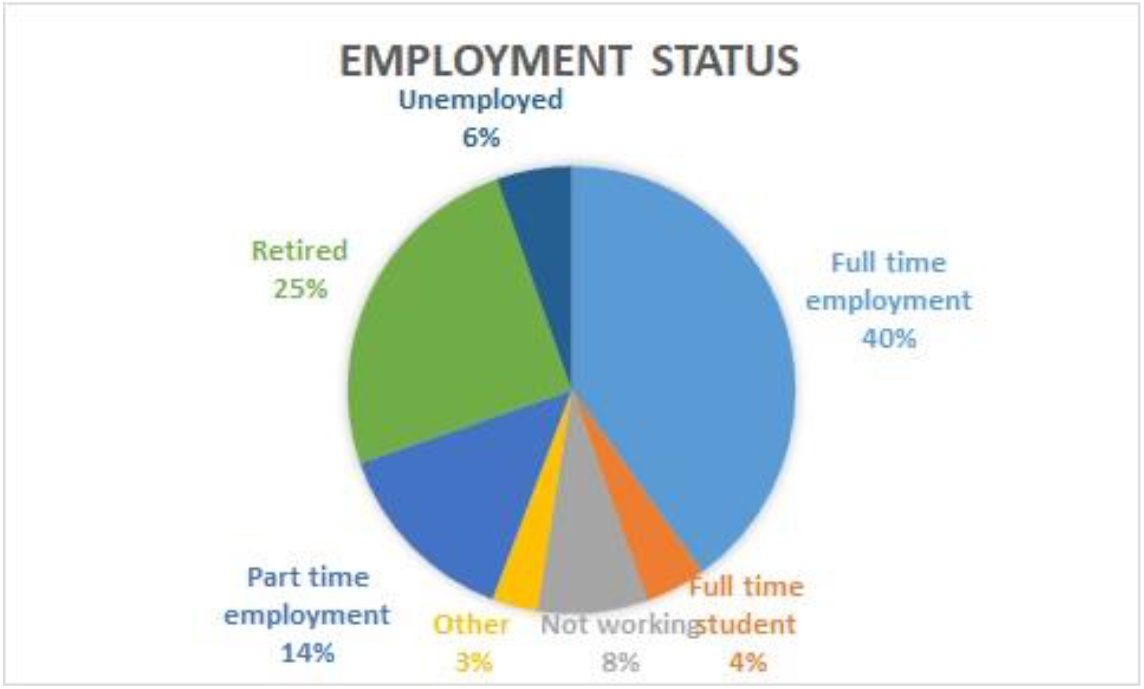
Employment Status.

Out of these, 2.7 % or 1131 individuals reported being carriers of COPD, whereas 19898 (47 %) stated that they did not have any of the 13 questioned conditions.

### Compliance

The share of COPD and healthy engaging in each of the behaviors are indicated in Figure 2.

**Figure 2.**
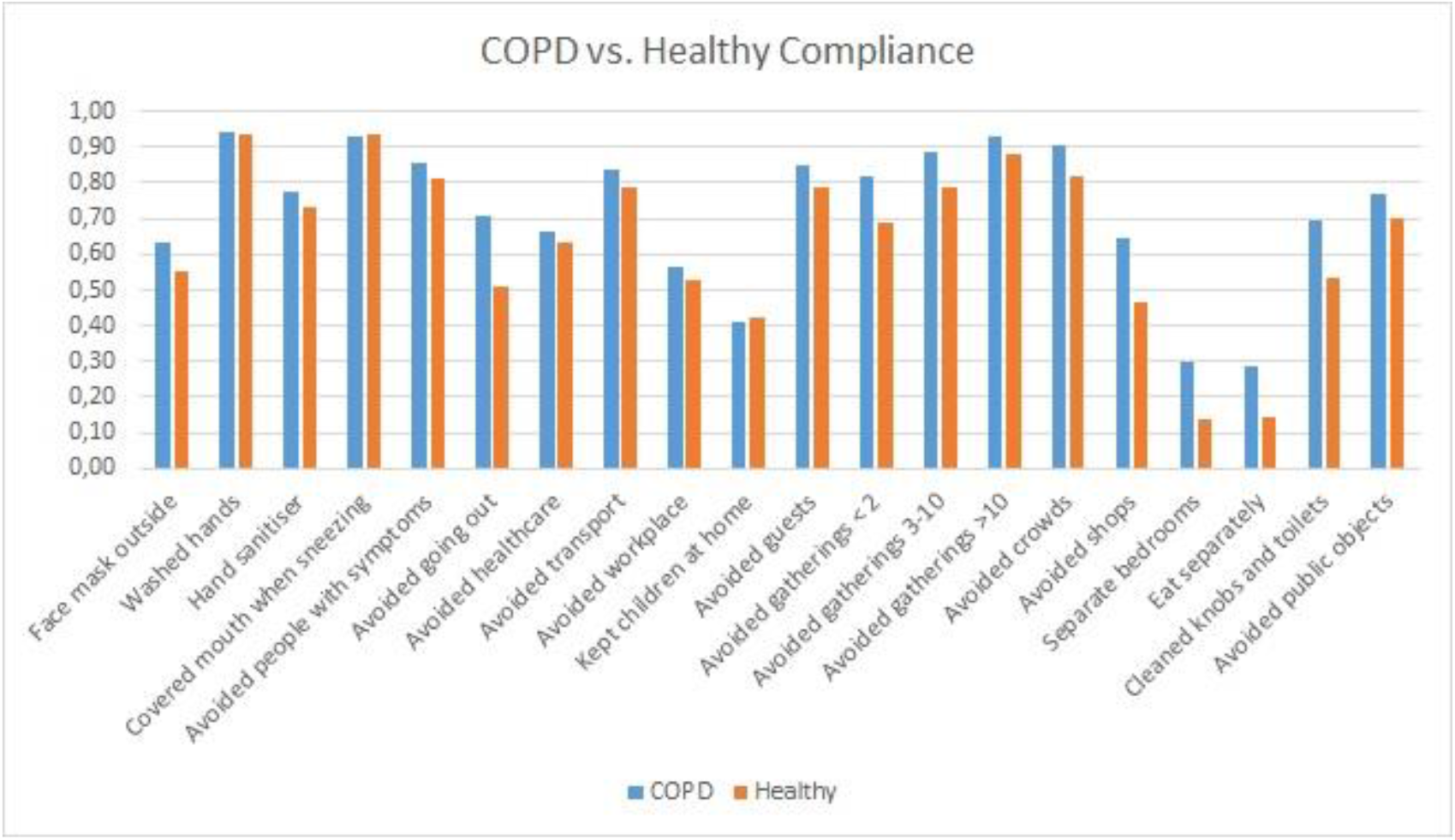
Social Distancing COPD vs. Healthy.

The results of hypothesis tests for proportions are summarised in Table 2.

**Table 2.** Hypothesis tests social distancing.

| Higher for COPD | No difference |
| --- | --- |
| Face mask outside | Washed hands |
| Hand sanitiser | Covered mouth when sneezing |
| Avoided people with symptoms | Kept children at home |
| Avoided going out |  |
| Avoided healthcare |  |
| Avoided transport |  |
| Avoided workplace |  |
| Avoided guests |  |
| Avoided gatherings < 2 |  |
| Avoided gatherings 3-10 |  |
| Avoided gatherings >10 |  |
| Avoided crowds |  |
| Avoided shops |  |
| Separate bedrooms |  |
| Eat separately |  |
| Cleaned knobs and toilets |  |
| Avoided public objects |  |

COPD patients were overall more stringent in nearly all categories.

### Time comparisons

Figure 3 illustrates the proportons for willingness to wear a face mask, showing the trends over time. Willingness to wear face coverings rose quickly in the spring of 2020. Those with COPD were more willing than non-COPD respondents especially during early phases of the pandemic, the share being higher for the first 5 months and then similar.

**Figure 3.**
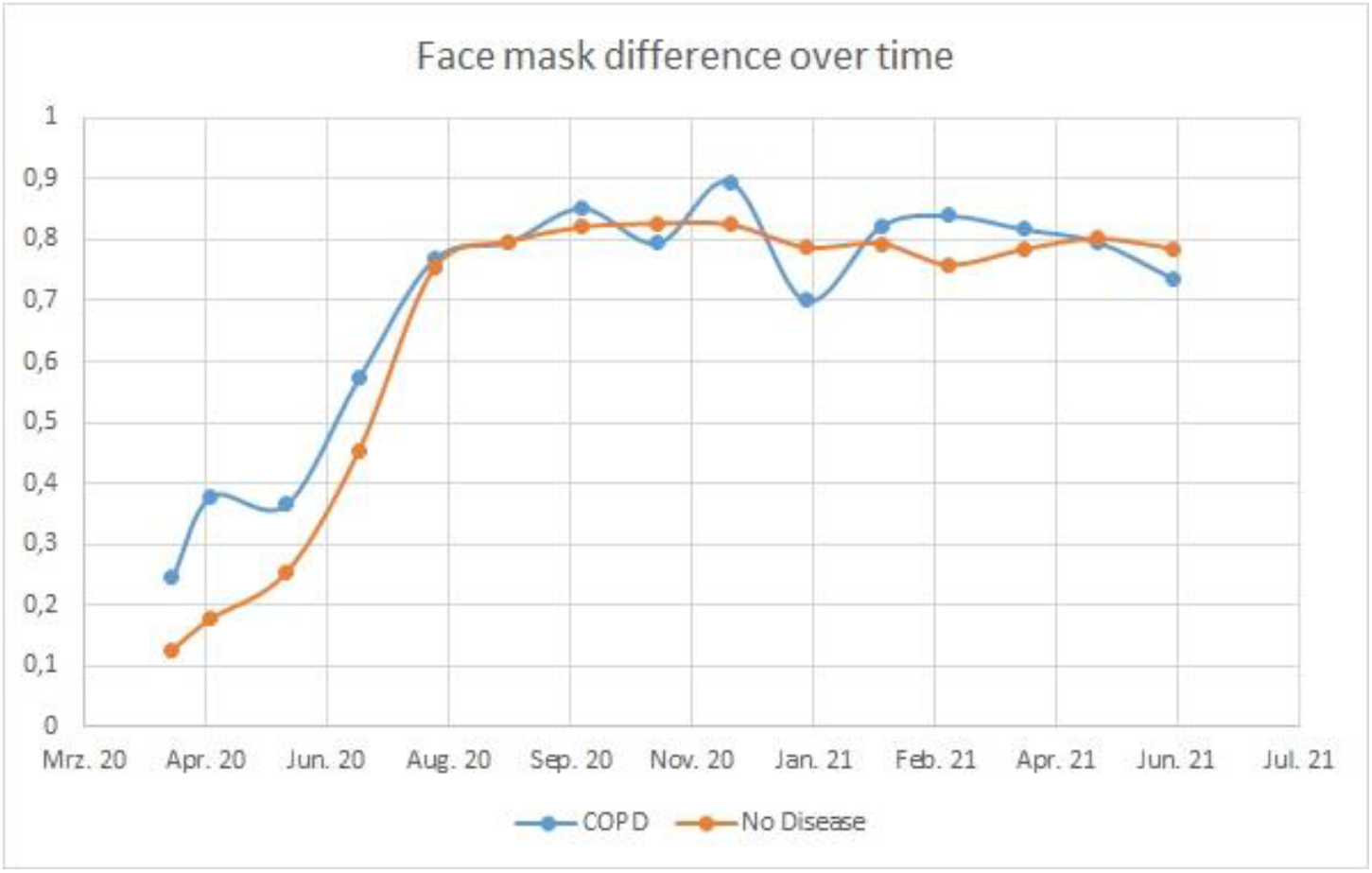
COPD and Face covering.

Consecutive hypothesis tests (Z test for proportions) revealed a significantly higher share in the COPD group from April to June 2020 and after that no significant differences.

Those in the COPD group indicated greater reluctance to going out in general throughout the period examined (Figure 4). This difference was significant for all months except May 2021.

**Figure 4.**
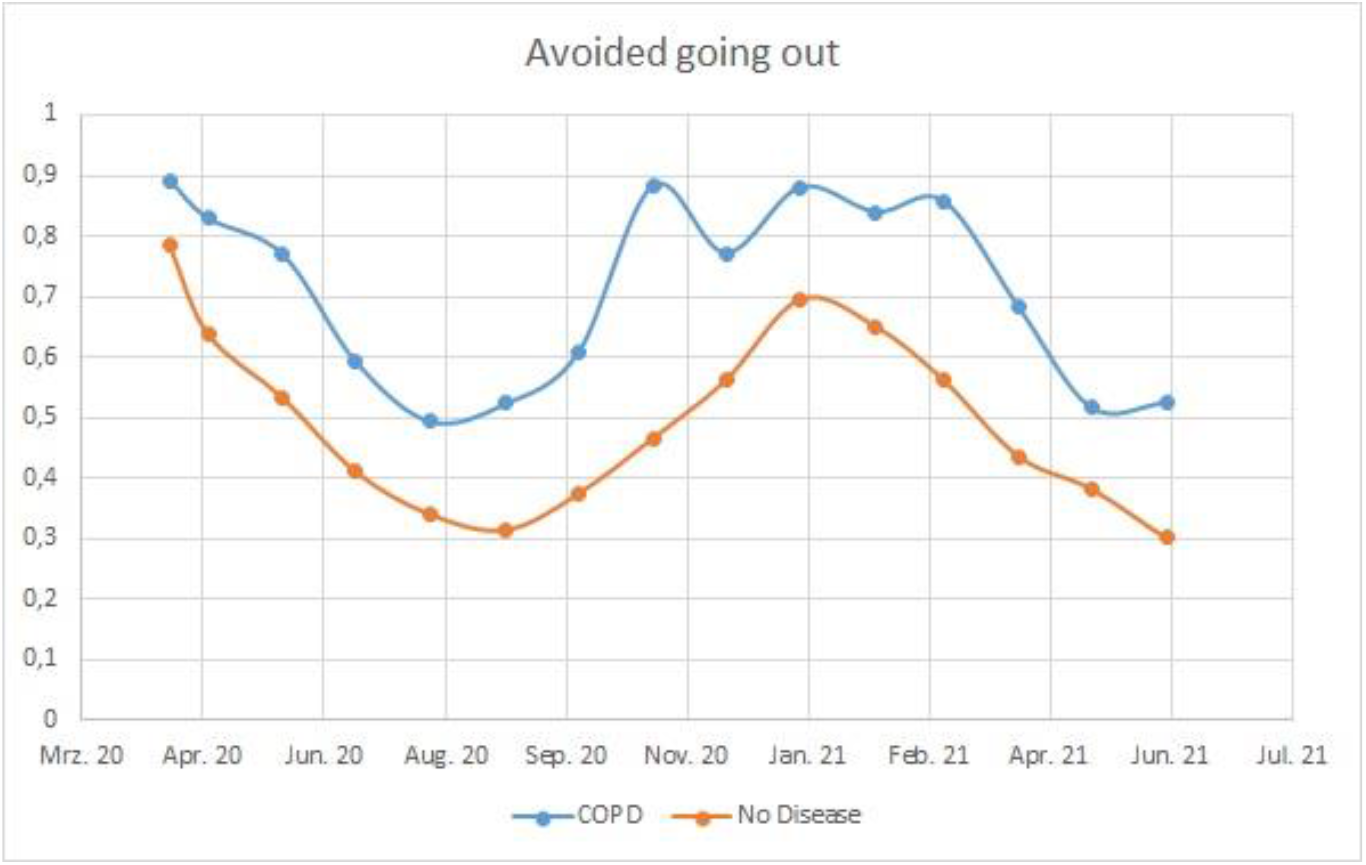
COPD and avoiding going outside.

In contrast, there was no apparent or significant difference in willingness to forego public transport usage between the groups (Figure 5). The hypothesis test showed significance only for October 2020.

**Figure 5.**
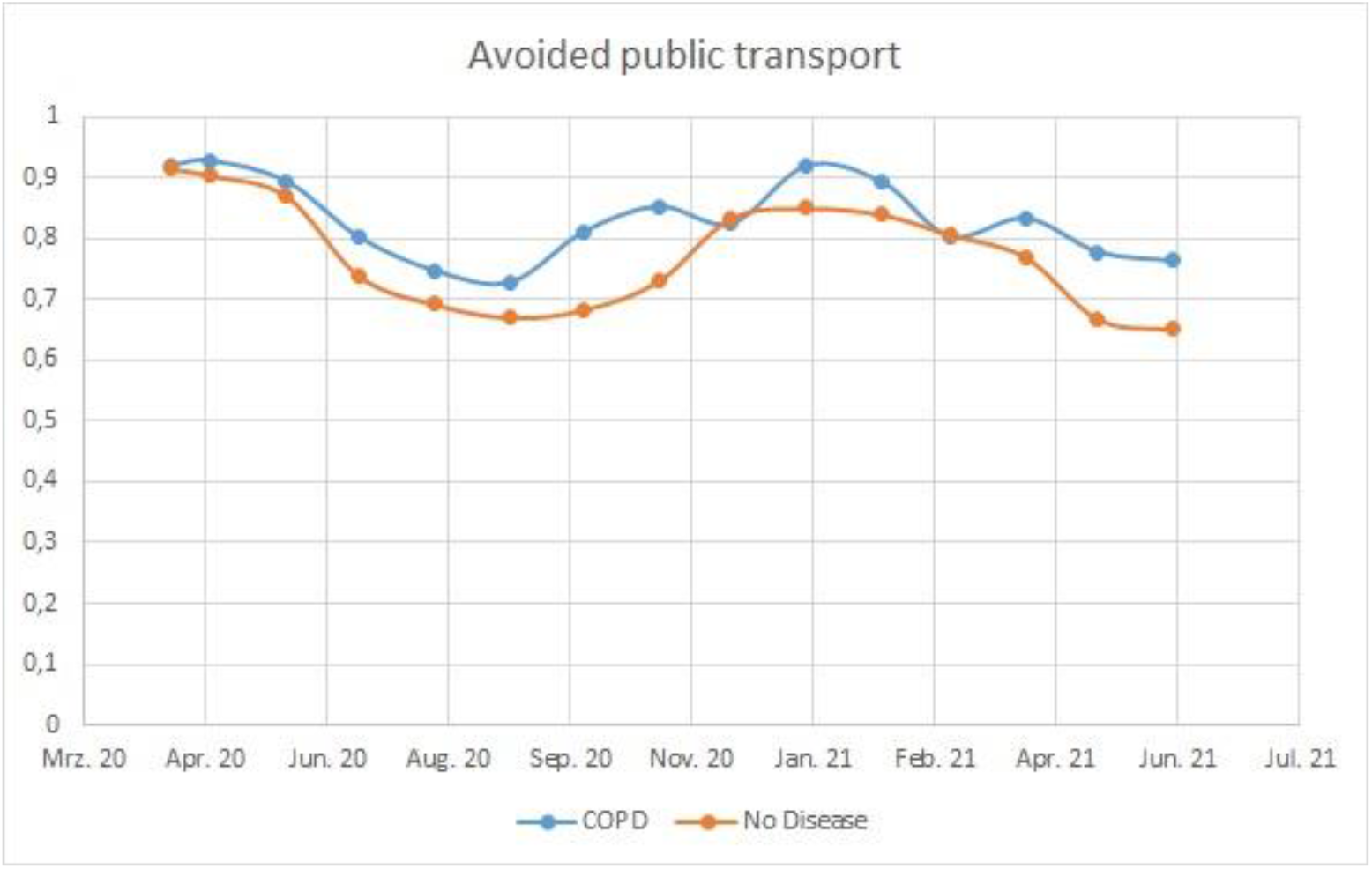
COPD and transport avoidance.

Similarly there was more reluctance to go shopping in the COPD groups through the entire period, as seen in Figure 6.

**Figure 6.**
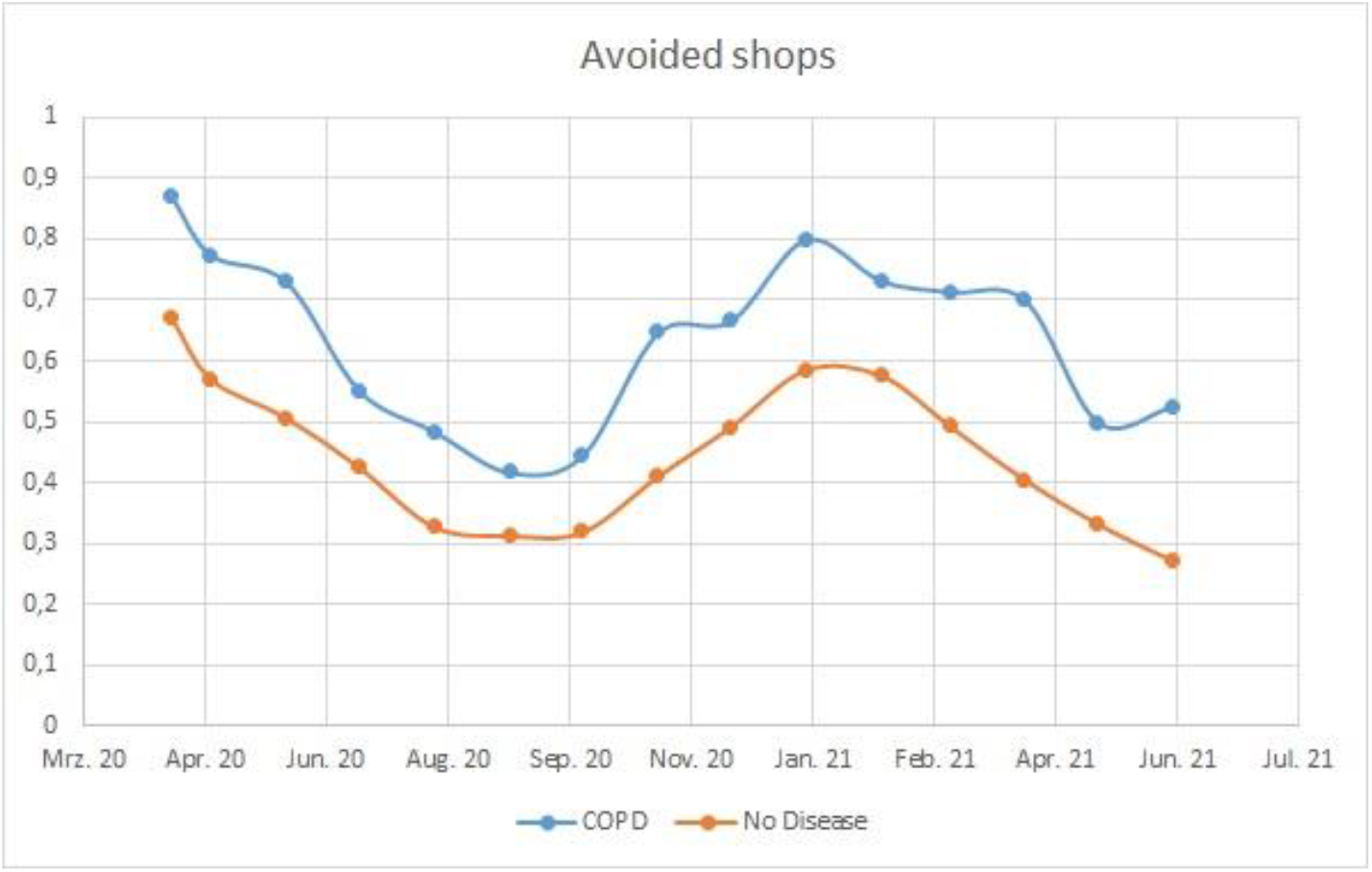
COPD and avoiding shops.

There was no apparent relationship between the number of COVID-19 cases in the week prior to surveying and levels of compliance to each question. Spearman Rank Correlations were weak for face mask wearing (COPD: 0.39, p = <0.01; non-COPD: 0.47, p = <0.01), avoiding going out (COPD: 0.2; non-COPD: 0.2), avoiding public transport (COPD:0.05, non-COPD:-0.04) and avoiding going shopping (COPD: 0.02; non-COPD: -0.04).

The mean age for COPD respondents was 65.18 (s = 12.05), while that of non-COPD respondents was 48.45 (s = 17.13). Results of a logistic regression comparing the influence of age, week and COPD status on answers showed that for face mask wearing COPD was the only significant variable (<0.01), with age and week of survey not being significant. For responses to going out and shopping questions, all variables were found to be significant. For the public transport question, age and week of survey were significant, COPD status was not.

## Discussion

### Summary

The results suggest that those with COPD were more willing to follow COVID-19 guidance and adhere to regulations than the general population for the large majority of recommended measures, especially those related to avoiding contact with other people and isolating socially. When looking in detail at compliance over time, it is clear that the difference was even larger during periods when the infection rate was at peak levels. While age was an important factor, logistic regression models indicated that having COPD was still a significant even when age was accounted. These patterns are to be expected intuitively, given that those with COPD would be at greater risk of experiencing complications following contraction of COVID-19 (1, 2). Although no relationship between case numbers and compliance was found, those with COPD may have perceived themselves as being at greater personal risk from COVID-19 due to the information they received on the condition.

### Strengths and limitations

The availability of the Imperial ‘COVID-19 Behavioural Tracker’ dataset offers the unique potential to examine attitudes of those with particular pre-existing health complaints, and whether they differ from the general population. A major benefit is the regular nature over which surveying was performed means that trends in adherence and attitudes could be examined over time. Most previous surveys are conducted at only a single point of time, providing only a ‘snapshot’ of attitudes with no relation to changing legislation or state of the Pandemic. The results show that such attitudes can vary over time. Examination of the factors underlying such attitude changes need to be examined.

Limitations are that only a single condition was considered here, COPD. Respondents were questioned as to a range of other health care conditions, including high blood pressure and cancer. Levels of adherence were not examined for other health problems, as a number of those, such as high blood pressure are unlikely to lead to elevated COVID-19 risk. Those with COPD are perceived to be, and indeed were, at great risk if they experienced COVID-19 infection. Another limitation is that underlying factors influencing adherence were not fully examined. The results of logistic regression determined that the presence of COPD was of most importance in face mask wearance. It was not significant in avoidance of public transport, as one might expect if this public transport use could not be avoided.

All presented results are based on self-reporting, and the the accuracy have been claimed to suffer from social desirability bias (16). However, a comprehensive review concluded that online self surveying results normally can be considered to be reliable (17).

### Comparison with existing literature

The factors determining adherence to health protection measures in general are complex and the subject of frequent study (8). Factors such as socio-economic status, gender, personal perceived level of risk, and quality of healthcare information all play a role. Examination of the factors potentially affecting behaviour during the COVID-19 pandemic has already begun, with studies identifying age and gender (9, 10) or income (11) as of importance. Another identified the perceived effectiveness of measures as to whether adherence was followed (12).

One study, where 2,000 adults were surveyed, examined those with pre-existing health problems found that are more likely to adhere to guidance (13). This found compliance to particular aspects varied depending on type of issue, for example those with cardiometabolic problems were more willing to remain home, while those with immune conditions were more willing to wear face coverings. In general people with pre-existing diseases were more willing to comply.

Specifically examining COPD, surveying of patients found they were more anxious than normal about their condition during the pre-lockdown phase, and that this increased to 58% as being more anxious during actual lockdown (14). Additionally 56% of COPD patients stated they avoided hospital during lockdown periods. The proportion of those where shopping was delivered or done by others rose from 23.8% to 70% between pre- and actual lockdown

The results presented here about face coverings may be surprising, as those with COPD could be excused for being particularly reluctant to wear such coverings, as they potentially hinder free breathing. Further, it is generally acknowledged that usage of facial coverings reduces the risk of transmission to others, rather than preventing contracting infection oneself. Differences in willingness to wear face masks declined over term, with the general population ‘catching up’ in terms of willingness compared to COPD sufferers. This may reflect legislation which mandated face mask use in the Health Protection (England) Regulations 2020, coming into force on 23 July 2020. In fact, there is some evidence that wearing face coverings has reduced the incidence of viral infections in general by those experiencing COPD resulting in a decline in COPD exacerbations (15).

### Implications for research and/or practice

Do patients comply when offered guidance and advice by GP’s or healthcare professionals? The results here indicate that such guidance could be important. Those with COPD exhibited more cautious behaviour than the general population when faced with COVID-19, probably due to the information they received which emphasised their risks. However, understanding the motivations behind such adherence is difficult. Personal perceptions of risk are important in determining adherence to health guidance. Those with COPD pursued greater self distancing measures, even before evidence that they were at greater risk of COVID-19 became available. Guidance provided by clinicians could well influence patient behaviours and have a positive effect on efforts to re-inforce cautious behaviours.

## Data Availability

Human data publicly available at the Imperial Behaviour Data Tracker:
https://github.com/YouGov-Data/covid-19-tracker

